# Longitudinal impact of different treatment sequences of second-generation antipsychotics on metabolic outcomes: a study using targeted maximum likelihood estimation

**DOI:** 10.1101/2024.10.25.24315949

**Authors:** Yaning Feng, Kenneth Chi-Yin Wong, Perry Bok-Man Leung, Benedict Ka-Wa Lee, Pak-Chung Sham, Simon Sai-Yu Lui, Hon-Cheong So

## Abstract

**Background:** Second-generation antipsychotics (SGAs) cause metabolic side-effects. However, patients’ metabolic profiles were influenced by many time-invariant and time-varying confounders. Real-world evidence on the long-term, dynamic effects of SGAs (e.g. different treatment sequences) is limited. We employed advanced causal inference methods to evaluate metabolic impact of SGAs in a naturalistic cohort.

**Methods:** We followed 696 Chinese patients with schizophrenia-spectrum disorders who received SGAs. Longitudinal targeted maximum likelihood estimation (LTMLE) was used to estimate the average treatment effects (ATEs) of continuous SGA treatment versus “no treatment” on metabolic outcomes, including total cholesterol (TC), high-density lipoprotein (HDL), low-density lipoprotein (LDL), triglyceride (TG), fasting glucose (FG), and body-mass-index (BMI), over 6-18 months at 3-month intervals. LTMLE accounted for both time-invariant and time-varying confounders. We also evaluated whether side-effects persisted after SGA discontinuation.

**Results:** The average treatment effects (ATEs) of continuous SGA treatment on BMI and TG showed an inverted U-shaped pattern, peaking at 12 months and declining afterwards. Similar patterns were observed for TC and LDL, albeit the ATEs peaked at 15 months. For FG and HDL, the ATEs peaked at ∼6 months. The adverse impact of SGAs on BMI persisted even after medication discontinuation, yet other metabolic parameters did not show such lingering side-effects. Compared with other SGAs, clozapine and olanzapine showed greater metabolic side-effects.

**Conclusions:** Our real-world study suggests that metabolic side-effects may stabilize with prolonged continuous treatment. Clozapine and olanzapine confer greater cardiometabolic risks than other SGAs. The side-effects of SGAs on BMI may persist after drug discontinuation. These insights may guide antipsychotic choice and help improve the management of metabolic side-effects.

## Introduction

Second-generation antipsychotics (SGAs) are commonly used as mainstream treatments for schizophrenia and related psychosis^1^, and have greater tolerability and fewer extrapyramidal side-effects than first-generation antipsychotics^2^. However, evidence from randomized controlled trials (RCTs) and population-based data indicated that SGAs are associated with metabolic side-effects, including weight gain, dyslipidemia, hyperglycemia and diabetes mellitus^3^, which warrants regular monitoring^4^ and psychoeducation^5^.

A previous systematic review^6^ of “head-to-head” comparisons of nine different SGAs revealed that these drugs differed in terms of the risk and severity of metabolic side-effects. More recently, a network meta-analysis^7^ pooled data from more than 100 RCTs (with a median of 6 weeks of follow-up) to compare and rank different antipsychotics according to their metabolic side-effects. The results revealed that olanzapine and clozapine had the worst side-effect profiles, whereas aripiprazole, brexpiprazole, cariprazine, lurasidone, and ziprasidone appeared more favorable in terms of metabolic side-effects. Another network meta-analysis^8^ of RCTs compared and ranked 31 first- and second-generation antipsychotics according to metabolic side-effects over a median of 45 weeks of follow-up, and revealed that olanzapine and clozapine were the two SGAs with the worst metabolic side-effects. Nevertheless, side-effect evaluation from longer follow-ups is lacking, and more real-world studies are warranted to improve generalizability of the findings.

Patients with schizophrenia and related psychosis usually receive long-term antipsychotic treatments, and clinicians may consider “phase-specific” care to address patients’ changing needs throughout the illness (e.g., the use of “minimum effective dose” of SGAs for stabilized remitted patients). Given that the efficacy, preparations and side-effects of SGAs differ considerably, “switching” between different antipsychotics is also common when patients develop extrapyramidal, metabolic or other (such as cardiac and gynecological) side-effects, treatment nonresponse, or problems in treatment adherence^9^. This highlights the *dynamic* nature of antipsychotic prescriptions in psychosis, which was very seldom investigated in previous studies.

Moreover, metabolic drugs such as metformin and simvastatin are commonly prescribed to patients who have developed more severe metabolic side-effects after receiving SGAs. These glucose- or lipid-lowering drugs can act as “time-varying confounders”, when the side-effects of SGAs are estimated using observational data. In addition, recent evidence^10^ suggests that the metabolic side-effects of SGAs can vary with combinations of other psychotropic drugs, prior side-effects, and patients’ current metabolite profiles. Although the design of RCT is robust, it cannot account for these “dynamic”, time-varying factors or confounders. Likewise, the two recent network meta-analyses^7^ of prior RCT data^8^ could not address the complex phenomenon of switching between SGAs and their subsequent cumulative effects on patients’ metabolic profiles. Besides, it is very difficult for RCTs to study how different “*sequences”* of treatments, which can vary over time, affect the severity of side-effects. As mentioned above, the median durations of follow-up in previous RCTs were relatively short (i.e., 6 weeks only in a recent network meta-analysis^7^). Moreover, many previous studies^7^ in this area only recruited Caucasians as subjects^8^, yet Chinese patients who received the same SGA may exhibit different patterns of metabolic changes^11^. It is therefore necessary to examine naturalistic and observational data in a cohort of patients who received SGAs for a longer term (preferably in non-Caucasian populations) to capture these real-world complexities.^7,8,11^

Time-varying confounders should be accounted for when estimating the causal effects of SGAs on metabolic side effects. These include metabolic drugs (like metformin and simvastatin), the *sequences* of SGA prescriptions, and patients’ (intermediate) metabolic status. However, conventional analytical methods^12^, including time-dependent Cox regression, random effects models, and generalized estimating equations, are known to produce biased estimates of causal effects in the presence of time-varying confounders^13^. Correcting for time-varying confounding factors requires more complex statistical methodologies^14^.

Targeted maximum likelihood estimation (TMLE)^14^ is a doubly robust method for estimating causal effects. Longitudinal TMLE (LTMLE)^15^ extends the principles of the TMLE to accommodate the complexities inherent in longitudinal studies, including time-varying treatments and confounders. Specifically, the LTMLE framework has the following advantages for studying the metabolic side-effects of SGAs:

1. Firstly, the framework can handle ***time-varying treatment***, which cannot be readily accounted for by other conventional methods. We wish to study in particular how a ***sequence of (dynamic) treatments***, which may change over time, affects metabolic parameters in patients. For example, assuming 3 time points of follow-up, a subject may be treated continuously all along (1,1,1) (1 indicating treatment), or just treated at the first time point (1,0,0), or during the first two time points (1,1,0) etc. To our knowledge, all previous studies on metabolic side-effects of SGAs only focused on treatment status defined cross-sectionally, instead of the whole sequence.
2. LTMLE can also handle ***time-varying confounders/covariates***, such as concomitant drugs. As such, LTMLE is considered an established method for causal inference in longitudinal studies due to its ability to tackle complex confounding patterns. Other commonly used methods, such as linear mixed models^16^, cannot readily handle time-varying covariates.
3. LTMLE is ***doubly robust***, meaning that it provides consistent estimates if either the outcome model or the treatment mechanism is correctly specified. This property gives LTMLE an advantage over other methods such as mixed models, which typically require correct specification of the outcome model. Double robustness also enhances the accuracy of causal effect estimates.

Given these advantages, we employed the LTMLE framework^17^ to study the “joint treatment effects” of different “SGA treatment sequences” on metabolic profiles, including total cholesterol (TC), high-density lipoprotein (HDL), low-density lipoprotein (LDL), triglyceride (TG), fasting glucose (FG) and body mass index (BMI) levels.

In summary, this study addressed the following objectives:

(1) To study the side-effects of SGAs under continuous treatment (defined by medication status every 1 or 3 months) for various durations (6, 9, 12, 15, and 18 months), compared to no treatment throughout the follow-up period; (2) To evaluate whether the side-effects of SGAs on metabolic parameters may persist despite discontinuation. To address this question, we compared metabolic outcomes at the end of 12 months, for SGAs taken for varying durations (e.g. 3, 6, 9, 12 months) versus no treatment all along; (3) To estimate the causal effects of SGAs on metabolic outcomes, accounting for time-varying confounding using a doubly robust framework. In addition, we also evaluated whether clozapine and olanzapine were causally linked to more severe metabolic side-effects compared to other SGAs.

Our approach allows for a more comprehensive and accurate assessment of the long-term metabolic effects of SGAs in real-world settings, accounting for the complexities of treatment patterns and time-varying confounders.

## Methods

### Our sample

We recruited 768 patients with schizophrenia spectrum disorders from the outpatient clinic at Castle Peak Hospital (CPH) in Hong Kong. The inclusion criteria were (1) Han Chinese ethnicity, (2) aged 18 years or above, (3) a diagnosis of schizophrenia or schizoaffective disorder according to the ICD-10^18^, (4) metabolic outcome measures (defined in the next section) available at three or more time points, and (5) at least three outcome measures available at a single time point. Patients were excluded if they had (1) a known history of metabolic disorders (e.g., diabetes mellitus or dyslipidemia) before SGA medications, or (2) lacked psychiatric follow-up as of March 2021. We retrieved the subjects’ electronic health records to collect detailed prescription history and outcome measures, including TC, HDL, LDL, TG, FG, and BMI, over the naturalistic follow-up period. In our analysis, the first prescription date was designated time 0. A total of 696 patients were included in the final analysis, with the age ranging from 19 to 73, and 54% of the sample was female.

### Outcome Variables

We captured six metabolic indicators as outcomes, namely, TC, HDL, LDL, TG, FG and BMI. As our dataset was based on a naturalistic cohort, participants’ metabolic profiles were not measured at the same time points^19^ (i.e., an unbalanced dataset). To convert the unbalanced dataset into a balanced dataset, we employed a linear mixed model (LMM)^20^ to impute the values of metabolic parameters at pre-specified time-points^18^. In brief, we included indicators of the drugs prescribed, treatment durations, and patients’ demographics (including age, sex, and education level) as predictors in our LMM, while records were grouped by patient ID with their own random intercept and slope. The methods are detailed in Supplementary Materials (Appendix A).

### Exposure Variables

The exposure variables in our study represented the use of different SGAs in our sample, including *clozapine, olanzapine, amisulpride, paliperidone, risperidone, quetiapine, and lurasidone* (Supplementary Table S1). We coded the exposure as a binary indicator, assigning a value of 1 if a subject had taken any of the seven SGAs at a given time point of interest and 0 if they had not (i.e., time point-based). For example, if a subject received any of these SGAs at time *t*, the exposure variable A_t_ was coded as 1; otherwise, it was coded as 0. Aripiprazole was excluded from the primary analysis because evidence from previous studies indicates that it is generally not associated with adverse metabolic outcomes^21^. However, to ensure the robustness of our findings, we included aripiprazole in the sensitivity analyses.

### Confounding variables

Both time-invariant and time-varying confounding variables were included in our longitudinal analysis. Patients’ age, sex, and baseline metabolic measures at *t*_O_ were included as baseline confounders. We coded several drugs, including metformin, atorvastatin, simvastatin and valproate, as “time-varying” confounders (Supplementary Table S1). Metformin, atorvastatin and simvastatin are metabolic drugs which lower glucose and lipid levels and can reduce weight. Valproate is a mood stabilizer associated with side-effect of weight gain and metabolic abnormalities^22,23^, and a substantial number of patients (*N*=82) in our sample received valproate. SGA prescription status across the follow-up period was also included by default as a time-varying covariate in the model (see explanation below). In addition, we included the mean age of each subject over their follow-up period as a time-invariant confounder.

Notably, by default, the LTMLE model for the dependent variables included all the “parent nodes” of the preceding time-points (see Appendix B of Supplementary Materials). **Fig. 1A** provides an illustration of the LTMLE model. For example, the outcome (*Y_3_*) at the 3^rd^ time-point (*t_3_*) was modeled on the basis of all the covariates/confounders (*L*), treatments (*A*) and outcomes (*Y*) at *all* previous time-points (*t_0_, t_1_, t_2_*). The inclusion of a wide range of covariates ensured proper control for time-varying confounding variables and provided a robust estimate of the causal effect.

**Fig. 1.**
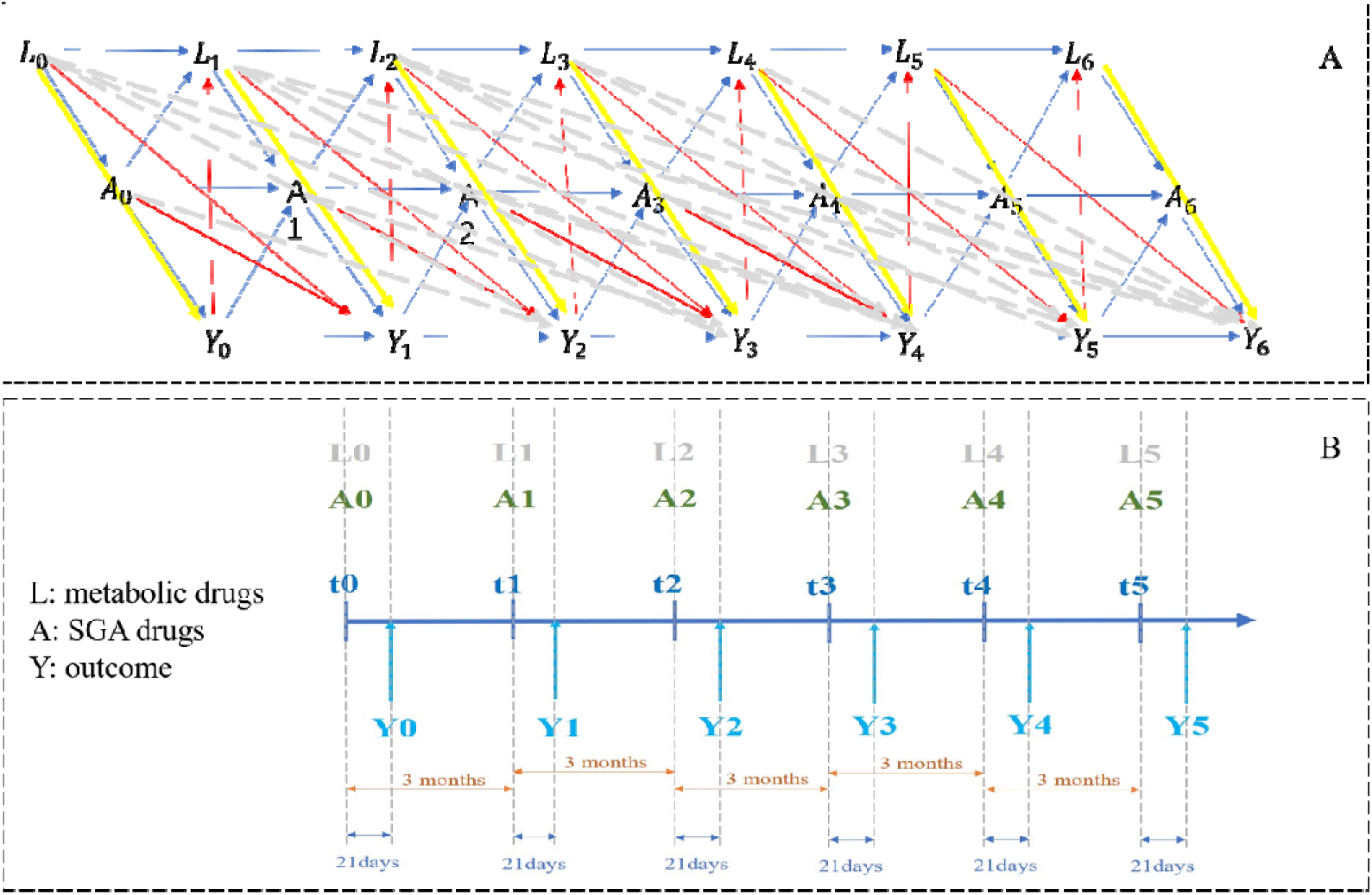
Assumed directed acyclic graph (A) and the sequential relationships among exposure (B), with the outcome and time-varying confounders at different time-points. L: Time-varying confounders A: Treatment at each time-point Y: Outcome at each time-point

### Statistical analysis using the LTMLE

We employed LTMLE^17^ to analyze the joint effects of taking SGAs on different metabolic indicators compared to those who have never taken SGAs. The LTMLE is a doubly robust method for estimating causal effects^24^, as it combines both an outcome model and a propensity score (PS)-based treatment model, which reduces bias from potential model misspecification. The TMLE methodology comprises three main steps: (1) using treatment and confounders as predictors to estimate the initial expected outcomes; (2) estimating the PS, which is the probability of treatment conditioned on covariates; and (3) using the PS to update the initial outcome model. This methodology is considered ‘doubly robust’ because it yields correct estimates if either the outcome model or the treatment mechanism is correctly specified. Details can be found in Appendix C of Supplementary Materials.

### Estimating the joint effect of treatment

Using the LTMLE framework, we estimated the metabolic effects of taking SGAs during the follow-up period, by comparing the metabolic profiles of patients treated with SGAs to those of patients never treated with SGAs. Specifically, we estimated the ***average treatment effects*** (ATEs) of SGAs on metabolic parameters in two scenarios: Situation A (“what if SGAs were taken throughout the follow-up period”) and Situation B (“what if no SGAs were taken during the follow-up period”). Additionally, we compared the ATEs of clozapine and olanzapine with the ATEs of other SGAs. Details of the mathematic estimation of the ATEs can be found in Appendix D of Supplementary Materials.

We conducted our analyses on the basis of a counterfactual framework, with the assumed network structure illustrated in Fig. 1A. In this framework, *L* represents time-varying confounders, *A* represents the exposure or treatment variable (SGAs in our study), and *Y* represents the outcome. The numbers following each variable indicate different time-points. Since metabolic outcomes can influence SGA prescriptions, we included intermediate *Y* variables in our model for adjustment (see Fig. 1A). The date of the first medication record was defined as time 0 (t_O_) for each patient, with a 3-month interval between time-points.

Both *L*_t_ and *A*_t_ were defined at each time-point *t*, whereas *Y*_t_ was recorded 21 days after *L*_t_ and *A*_t_. We allowed this 21-day window because metabolic side effects may take time to develop; this window length was determined based on goodness-of-fit testing from our previous study^25^. Fig. 1B illustrates the sequential relationships between exposure, outcome and time-varying confounders at different time-points. We conducted these analyses via the R package ‘ltmle’^12^ (version 1.2.0).

### Sensitivity analyses

We conducted several sensitivity analyses to ensure the robustness of our findings. *First*, we removed the ‘intermediate outcomes’ from our model, which included the metabolic parameters at intermediate time-points, and we compared the results with and without this adjustment. This method allowed us to determine whether using a simpler model would affect the conclusions. *Second*, we set two different time intervals (1 and 3 months) while maintaining the same total follow-up period. *Third*, although previous studies have generally shown that aripiprazole seldom causes metabolic issues^7^, we included this SGA in our analysis to validate our findings further. *Fourth*, we addressed the possibility that patients might discontinue SGAs during the follow-up period by implementing an alternative exposure definition. Specifically, we defined exposure based on whether a patient received SGAs continuously during the observed time interval (i.e., interval-based). We set different cut-off values to determine whether exposure at time *t* should be coded as 1 or 0. For example, if the cut-off value is set as 0.5, we coded the exposure at time *t* as 1 if the patient was treated with SGAs for more than 50% of the time in the observed interval (*t, t*+1) (see Table S2).

## Results

### Target maximum likelihood estimation

#### Continuous SGA treatment vs no treatment (for different follow-up periods)

We first compared “what if all the participants were always treated during the follow-up period” versus “what if all the participants were never treated”. The results are shown in Fig. 1B and Table 1. We applied LTMLE to study the effects of continuous SGA treatment for follow-up durations of 6, 9, 12, 15, and 18 months.

**Table 1.**
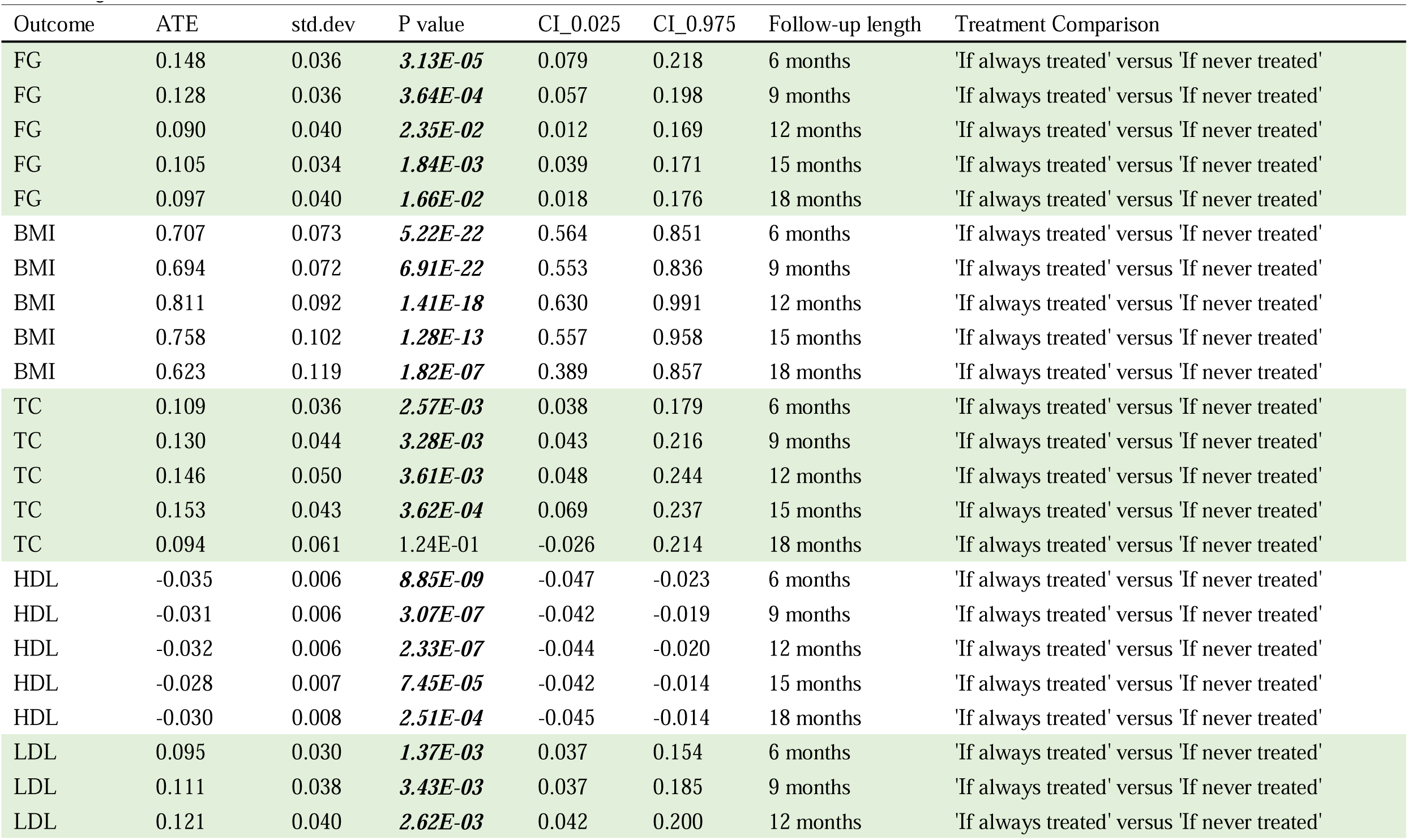
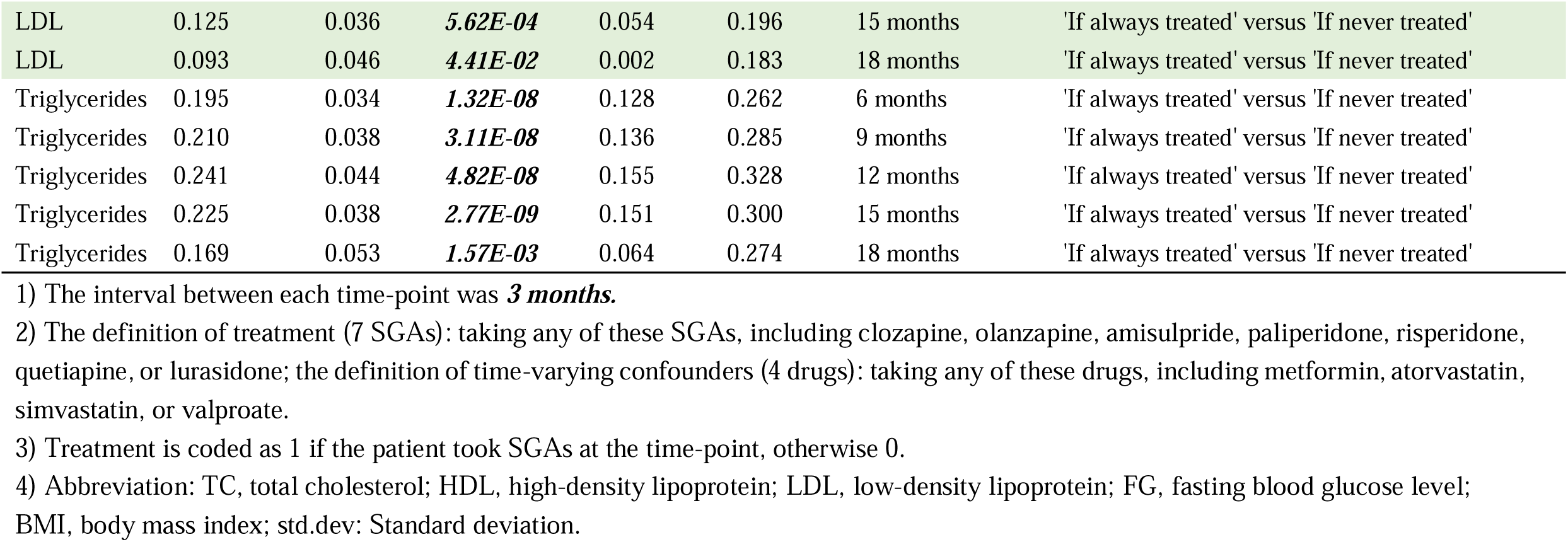
Average treatment effect (ATE) between ‘always being treated by SGAs’ and ‘never being treated by SGAs’ (with adjustment of intermediate outcomes)

| Outcome | ATE | std.dev | P value | CI_0.025 | CI_0.975 | Follow-up length | Treatment Comparison |
| --- | --- | --- | --- | --- | --- | --- | --- |
| FG | 0.148 | 0.036 | **3.13E-05** | 0.079 | 0.218 | 6 months | 'If always treated' versus 'If never treated' |
| FG | 0.128 | 0.036 | **3.64E-04** | 0.057 | 0.198 | 9 months | 'If always treated' versus 'If never treated' |
| FG | 0.090 | 0.040 | **2.35E-02** | 0.012 | 0.169 | 12 months | 'If always treated' versus 'If never treated' |
| FG | 0.105 | 0.034 | **1.84E-03** | 0.039 | 0.171 | 15 months | 'If always treated' versus 'If never treated' |
| FG | 0.097 | 0.040 | **1.66E-02** | 0.018 | 0.176 | 18 months | 'If always treated' versus 'If never treated' |
| BMI | 0.707 | 0.073 | **5.22E-22** | 0.564 | 0.851 | 6 months | 'If always treated' versus 'If never treated' |
| BMI | 0.694 | 0.072 | **6.91E-22** | 0.553 | 0.836 | 9 months | 'If always treated' versus 'If never treated' |
| BMI | 0.811 | 0.092 | **1.41E-18** | 0.630 | 0.991 | 12 months | 'If always treated' versus 'If never treated' |
| BMI | 0.758 | 0.102 | **1.28E-13** | 0.557 | 0.958 | 15 months | 'If always treated' versus 'If never treated' |
| BMI | 0.623 | 0.119 | **1.82E-07** | 0.389 | 0.857 | 18 months | 'If always treated' versus 'If never treated' |
| TC | 0.109 | 0.036 | **2.57E-03** | 0.038 | 0.179 | 6 months | 'If always treated' versus 'If never treated' |
| TC | 0.130 | 0.044 | **3.28E-03** | 0.043 | 0.216 | 9 months | 'If always treated' versus 'If never treated' |
| TC | 0.146 | 0.050 | **3.61E-03** | 0.048 | 0.244 | 12 months | 'If always treated' versus 'If never treated' |
| TC | 0.153 | 0.043 | **3.62E-04** | 0.069 | 0.237 | 15 months | 'If always treated' versus 'If never treated' |
| TC | 0.094 | 0.061 | 1.24E-01 | -0.026 | 0.214 | 18 months | 'If always treated' versus 'If never treated' |
| HDL | -0.035 | 0.006 | **8.85E-09** | -0.047 | -0.023 | 6 months | 'If always treated' versus 'If never treated' |
| HDL | -0.031 | 0.006 | **3.07E-07** | -0.042 | -0.019 | 9 months | 'If always treated' versus 'If never treated' |
| HDL | -0.032 | 0.006 | **2.33E-07** | -0.044 | -0.020 | 12 months | 'If always treated' versus 'If never treated' |
| HDL | -0.028 | 0.007 | **7.45E-05** | -0.042 | -0.014 | 15 months | 'If always treated' versus 'If never treated' |
| HDL | -0.030 | 0.008 | **2.51E-04** | -0.045 | -0.014 | 18 months | 'If always treated' versus 'If never treated' |
| LDL | 0.095 | 0.030 | **1.37E-03** | 0.037 | 0.154 | 6 months | 'If always treated' versus 'If never treated' |
| LDL | 0.111 | 0.038 | **3.43E-03** | 0.037 | 0.185 | 9 months | 'If always treated' versus 'If never treated' |
| LDL | 0.121 | 0.040 | **2.62E-03** | 0.042 | 0.200 | 12 months | 'If always treated' versus 'If never treated' |
| LDL | 0.125 | 0.036 | **5.62E-04** | 0.054 | 0.196 | 15 months | 'If always treated' versus 'If never treated' |
| LDL | 0.093 | 0.046 | **4.41E-02** | 0.002 | 0.183 | 18 months | 'If always treated' versus 'If never treated' |
| Triglycerides | 0.195 | 0.034 | **1.32E-08** | 0.128 | 0.262 | 6 months | 'If always treated' versus 'If never treated' |
| Triglycerides | 0.210 | 0.038 | **3.11E-08** | 0.136 | 0.285 | 9 months | 'If always treated' versus 'If never treated' |
| Triglycerides | 0.241 | 0.044 | **4.82E-08** | 0.155 | 0.328 | 12 months | 'If always treated' versus 'If never treated' |
| Triglycerides | 0.225 | 0.038 | **2.77E-09** | 0.151 | 0.300 | 15 months | 'If always treated' versus 'If never treated' |
| Triglycerides | 0.169 | 0.053 | **1.57E-03** | 0.064 | 0.274 | 18 months | 'If always treated' versus 'If never treated' |

1) The interval between each time-point was **3 months**.
2) The definition of treatment (7 SGAs): taking any of these SGAs, including clozapine, olanzapine, amisulpride, paliperidone, risperidone, quetiapine, or lurasidone; the definition of time-varying confounders (4 drugs): taking any of these drugs, including metformin, atorvastatin, simvastatin, or valproate.
3) Treatment is coded as 1 if the patient took SGAs at the time-point, otherwise 0.
4) Abbreviation: TC, total cholesterol; HDL, high-density lipoprotein; LDL, low-density lipoprotein; FG, fasting blood glucose level; BMI, body mass index; std.dev: Standard deviation.

Overall, for BMI and TG, ATEs initially increased and then decreased over time, when comparing ’always-treated’ to ’never-treated.’ A quadratic fit showed ATEs peaked at around 12 months for BMI and TG, and 15 months for TC and LDL. HDL fluctuated but increased from 6 to 18 months.

Specifically, positive ATEs were observed for BMI. At the 6-month follow-up, the ATE was 0.707 (95% CI = 0.564–0.851) kg/m², indicating that the BMI would be 0.707 kg/m² higher if SGAs were taken continuously during the follow-up period than if SGA treatment was not given. The ATE for BMI increased to 0.811 (95% CI = 0.63–0.991) kg/m² at 12 months and decreased to 0.623 (95% CI = 0.389–0.857) kg/m² at 18 months.

TG showed a similar pattern to that of BMI, with ATE increasing from 0.195 (95% CI = 0.128–0.262) mmol/L to 0.241 (95% CI = 0.155–0.328) mmol/L when the follow-up length increased from 6 months to 12 months and then decreased to 0.169 (95% CI = 0.064–0.274) mmol/L at 18 months.

The ATEs peaked at 15 months for TC and LDL. The ATE for TC increased from 0.109 (95% CI = 0.038–0.179) mmol/L at 6 months to 0.153 (95% CI = 0.069–0.237) mmol/L at 15 months, and then decreased to 0.094 (95% CI = -0.026–0.214) mmol/L at 18 months. The ATE for LDL increased from 0.095 (95% CI = 0.037–0.154) at 6 months to 0.125 (95% CI = 0.054–0.196) mmol/L at 15 months.

HDL showed no clear pattern, with slight fluctuations of ATEs over time. The most negative ATE for HDL occurred at 6 months, but it increased slightly from 6 to 15 months. For FG, the highest ATE was observed at 6 months, followed by a decreasing trend.

#### Sensitivity analysis with a 1-month interval

We performed sensitivity analysis by considering one-month follow-up intervals (ATEs evaluated from the 4^th^ month onward). As shown in Fig. 2, at 3-month intervals, the ATEs of most metabolic outcomes initially increased but then decreased. With 1-month intervals and the same follow-up periods, BMI, TG, TC, FG, and LDL showed similar patterns (Fig. S1), indicating that the observed trends were likely robust. The trends before 6 months are not captured in Fig. 2 but can be observed in Fig. S1.

**Fig. 2.**
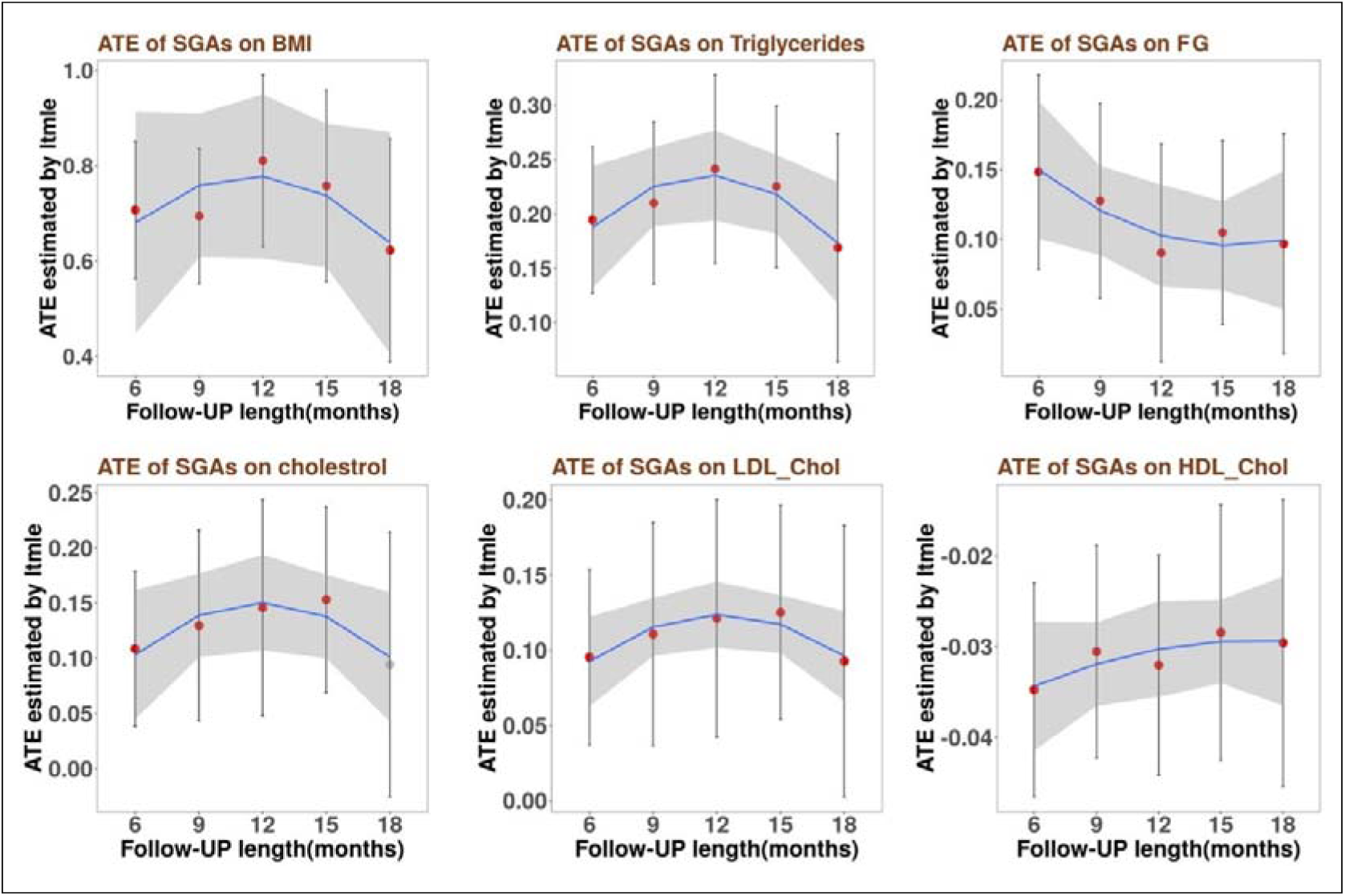
Average treatment effects (ATEs) of six outcomes for different FU lengths based on a 3-month interval. The red data points indicate a statistically significant difference (p < 0.05) in the outcome measure between ‘always being treated’ and ‘never being treated’ groups, whereas the gray data points represent nonsignificant differences. The blue line is generated on the basis of ‘ATE ∼ follow-up lengths + square (follow-up lengths)’, and the gray area indicates the 95% confidence interval. Abbreviations: TC, total cholesterol; HDL, high-density lipoprotein; LDL, low-density lipoprotein; FG, fasting blood glucose level; BMI, body mass index; FU, Follow-up.

Notably, for FG, the ATEs peaked at 4 months and then gradually decreased until 12 months, with a slightly increasing trend between 12 and 18 months (Fig. S1). The ATEs for HDL decreased between 4 and 6 months, followed by a relatively steady (but slightly increasing) trend from 6 to 18 months. These findings suggested that the side-effects of SGAs on HDL levels were the most pronounced at around 6 months after continuous treatment.

#### Alternative “interval-based” treatment definitions

With three additional *interval-based* treatment definitions (i.e., requiring treatment at least a certain proportion of time during the interval; see Table S2), similar patterns were observed, but with smaller ATEs (Table S5). For example, at 18 months, the ATE of SGAs on BMI was 0.623 (95% CI = 0.389-0.857) kg/m^2^ (Table 1) when we defined the treatment as “patients taking SGAs at the specified time points”, but it became 0.357 (95% CI = 0.136-0.579) kg/m² (Table S5) when we defined the treatment as “patients taking SGAs more than 80% of the time in the observed interval”.

#### Clozapine and olanzapine versus other SGAs

Given that clozapine and olanzapine are associated more serious metabolic side-effects in previous studies^8^, we stratified the groups of patients who started with SGAs and compared their differences between Situation A (“what if everyone in our sample took clozapine or olanzapine throughout the follow-up”) and Situation B (“what if everyone in our sample were on other SGAs during the follow-up”).

Table 3 shows the results of ATEs in these analyses. For example, a positive ATE was observed for BMI (ATE= 0.947 kg/m² (95% CI = 0.461-1.433) at 18 months). This indicates greater increase of BMI in patients treated with clozapine or olanzapine than in those treated with other SGAs after 18 months. Positive ATEs were also observed for FG, TC, LDL, and TG, with ATEs of 0.178 (95% CI = 0.067-0.29) mmol/L, 0.283 (95% CI = 0.118-0.447) mmol/L, 0.205 (95% CI = 0.083-0.327) mmol/L, and 0.36 (95% CI = 0.238-0.481) mmol/L, respectively. A negative ATE was observed for HDL (-0.064 mmol/L, 95% CI = -0.099-0.029), which suggested that patients treated with clozapine or olanzapine had 0.064 mmol/L lower HDL on average after 18 months.

#### Metabolic outcomes at 12 months with different durations of SGA treatment

We also compared the metabolic outcomes at 12 months, when “SGAs were taken for varying durations (e.g., 3, 6, 9, and 12 months)” versus “no treatment all along”. The results are shown in Table 3 and Fig. S2.

Interestingly, our findings suggested a possible “accumulative” or lingering side-effect of SGA treatment on BMI. Regardless of the duration of SGA treatment, BMI remained significantly elevated at 12 months compared with “no treatment”, even after the discontinuation of SGA. In contrast, this kind of lingering effect was not observed for other metabolic outcomes. Compared with “no SGA treatment all along”, the effect sizes were non-significant for all other metabolic outcomes (except for BMI), if the drug had been discontinued for at least 3 months before the final assessment date.

As an alternative approach to analyzing the cumulative effects of SGAs, we also compared SGAs taken for different durations with those under continuous treatment throughout the follow-up period (Table S4). If the SGA side-effects do not persist after discontinuation, we would expect the metabolic outcomes for SGA treatment durations less than the full follow-up period to be significantly better than those with SGAs taken all along. As shown in Table S4, we observed such patterns for almost all the metabolic outcomes.

With respect to BMI, we found that SGA discontinuation before the end of follow-up was associated with a significantly lower BMI than continued SGA treatment. In our previous analysis, we observed that BMI remained elevated despite discontinuation, but this was in comparison to those never treated with SGAs. Taken together, the findings suggest that for those who take SGAs for a limited duration during FU, their BMI might fall between those who are never treated with SGAs and those who receive continuous SGA treatment.

### Additional sensitivity analyses

First, we conducted a sensitivity analysis with a 1-month follow-up interval, as detailed above. Second, we compared the results with and without adjustment for the intermediate metabolic outcome values (Table 1 and Table S3). After such sensitivity analysis, we observed similar patterns of results.

Third, we included aripiprazole as an SGA and compared the ATEs (Table 1 and Table S6). The patterns of results remained almost the same, except that we observed slightly larger ATEs when aripiprazole was excluded, indicating a relatively small impact of aripiprazole on metabolic outcomes.

Fourth, we employed the interval-based treatment definition. When the results in Table 1 and Table S5 were compared, similar patterns of ATEs were observed, but ATEs were generally smaller. For example, at 12 months, the ATE of SGAs on BMI was 0.811 kg/m² (95% CI = 0.63-0.991) in Table 1, where treatment was defined as “patients taking SGAs at specific time-points,” but 0.379 kg/m² (95% CI = 0.196-0.562) in Table S5, where treatment was defined as “patients taking SGAs more than 80% of the time in the observed time interval”. It is reasonable that using a stricter criterion to define treatment would result in a smaller detectable difference between the ’treated’ and the ’untreated’ groups. Nonetheless, their difference (ATEs) remains significant, supporting the robustness of our findings.

## Discussion

In this study, we applied a longitudinal TMLE framework to study the joint effect of SGAs on metabolic indicators including TC, HDL, LDL, BMI, triglycerides, and FG. We estimated the ATE of being treated with SGAs for 6, 9, 12, 15, and 18 months. In addition, we studied the impact of treatment discontinuation on different outcomes. Sensitivity analyses were performed to validate the results.

### Main findings

In general, as follow-up length increased from 6 to 18 months, we observed increasing ATEs for BMI and TG in 6-12 months, followed by decreasing trends from 12 to 18 months. A similar pattern was noted for TC and LDL, although the peak ATEs were observed at 15 months instead. In other words, while the side-effects would initially increase after the commencement of SGAs, they would not escalate for a long time. Such findings aligned with previous meta-analyses which reported that antipsychotic-induced side-effects on lipids would develop rapidly but then stabilize ^9^.

Notably, ATEs typically peaked around 12–15 months and then slightly decreased. This pattern may be due to patients taking measures to counteract side-effects, such as diet modifications and more physical activities, which were not captured in our naturalistic dataset. It is also possible that patients developed “resilience” to the antipsychotic side-effects with prolonged treatment, though the underlying mechanisms will warrant further studies. On the other hand, we observed a different pattern for HDL and FG. For HDL, fluctuations were observed, with a clearer pattern of decrease followed by an increase in ATE emerging at 1-month intervals. This may be due to more frequent observations providing additional information over the same follow-up period. For FG, the trend differed from that of other metabolic parameters; however, we still observed significantly positive ATEs across different follow-up time points, indicating adverse effects of SGAs on FG. However, the increase of FG plateaued earlier than the other metabolic outcomes.

Notably, a lingering effect of SGAs on BMI was observed (Table 3 and Fig. S2). BMI remained elevated across different treatment sequences (from 1000 to 1111, i.e., four time-points with 3-month intervals) compared with no treatment (i.e., 0000), despite the discontinuation of SGAs before the end of follow-up.

Relatively few studies have examined the long-term (>1 year) effects of SGAs on a comprehensive panel of metabolic parameters in schizophrenia patients. Recently, Vázquez-Bourgon et al. performed a 10-year follow-up of first-episode psychosis patients and controls, and found that discontinuation of antipsychotics was associated with better metabolic profiles compared to continuous treatment, including less weight gain^26^. However, the metabolic profiles of patients who discontinued treatment were still worse than those of healthy controls. These findings suggested that metabolic side-effects may persist despite discontinuation of SGAs. In this study, we primarily found BMI to be persistently affected, contrary to Vázquez-Bourgon et al.’s findings that other metabolic parameters (e.g., HDL, TG and insulin resistance) were persistently affected. However, Vázquez-Bourgon et al. compared treatment discontinuers with healthy controls instead of other psychosis subjects, making it difficult to isolate the specific effects of SGAs from the metabolic impacts of psychosis itself. In another study, Mackin et al.^27^ compared patients who discontinued SGAs to those receiving continuous treatments, and reported that BMI and waist circumference increased in both groups, with no significant difference over a follow-up of four years. Besides, they did not find any significant difference in glucose and lipid measures. However, Mackin et al.’s study ^27^ had a small sample size (i.e., 89 subjects only). Taken together, the current and prior studies provided evidence that the metabolic effects of SGAs may persist to varying degrees, even after medication discontinuation, though more research with larger samples is needed. The persistence of weight gain after discontinuation of SGAs seemed to be a consistent finding, although other metabolic outcomes showed mixed results across studies. Finally, consistent with earlier findings^28^, we also observed larger ATEs when comparing clozapine/olanzapine with other SGAs (Table 2).

**Table 2.**
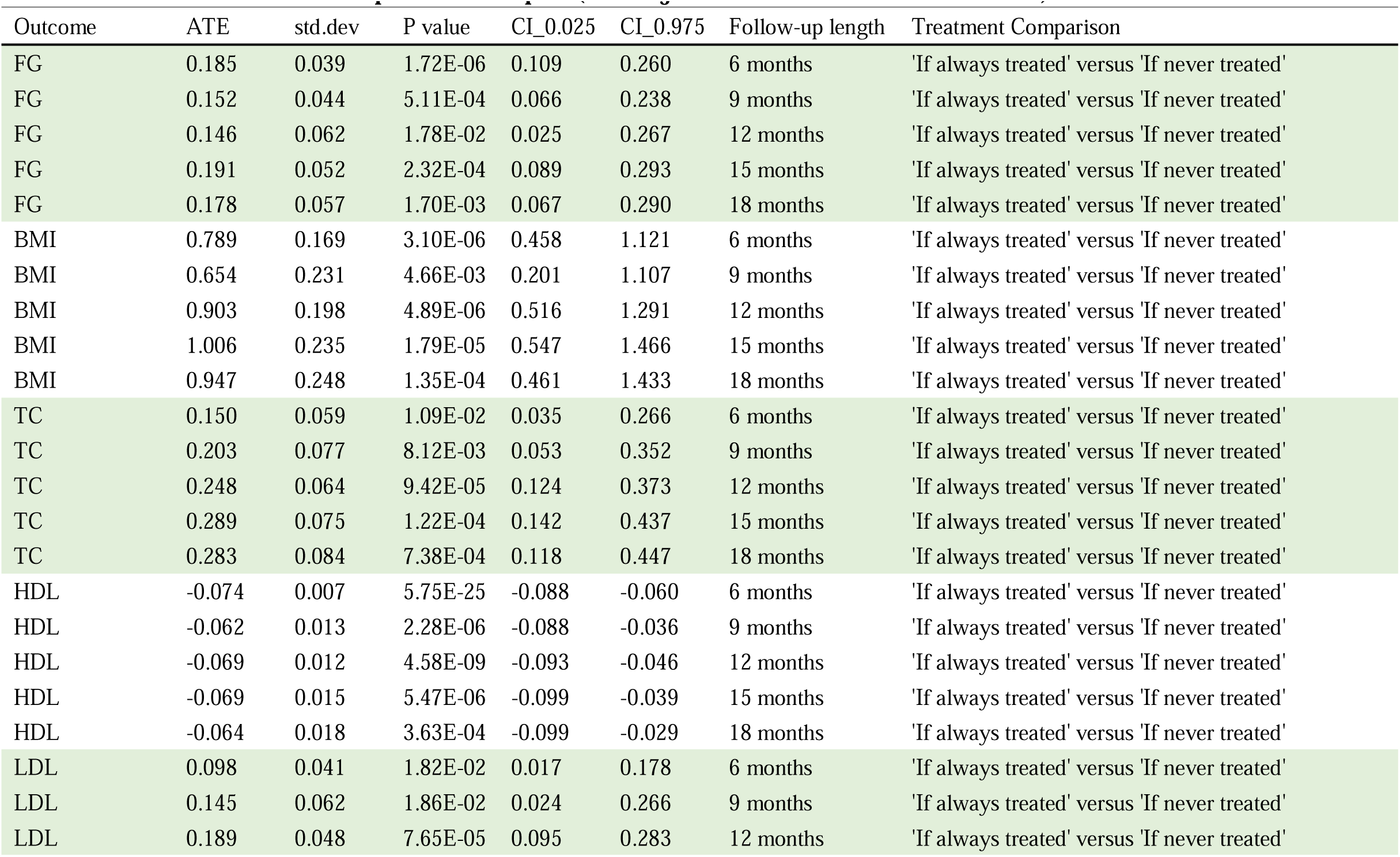
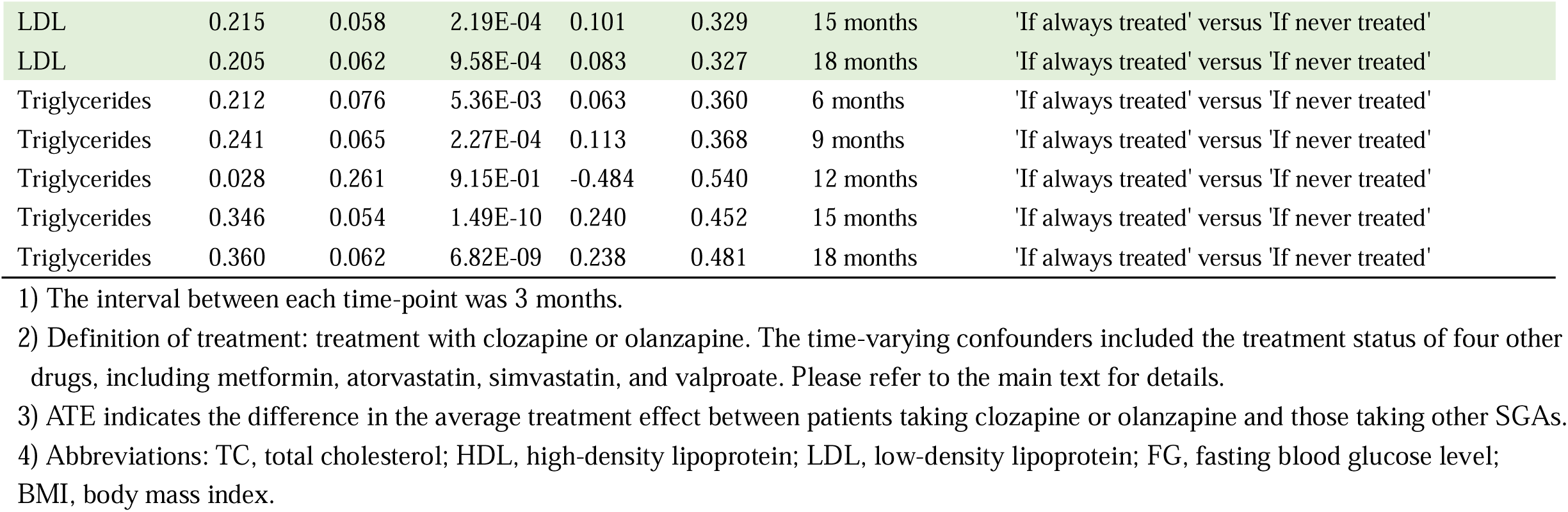
Average treatment effect (ATE) between patients treated with clozapine or olanzapine throughout the follow-up period and those never treated with clozapine or olanzapine (with adjustment for intermediate outcomes)

| Outcome | ATE | std.dev | P value | CI_0.025 | CI_0.975 | Follow-up length | Treatment Comparison |
| --- | --- | --- | --- | --- | --- | --- | --- |
| FG | 0.185 | 0.039 | 1.72E-06 | 0.109 | 0.260 | 6 months | 'If always treated' versus 'If never treated' |
| FG | 0.152 | 0.044 | 5.11E-04 | 0.066 | 0.238 | 9 months | 'If always treated' versus 'If never treated' |
| FG | 0.146 | 0.062 | 1.78E-02 | 0.025 | 0.267 | 12 months | 'If always treated' versus 'If never treated' |
| FG | 0.191 | 0.052 | 2.32E-04 | 0.089 | 0.293 | 15 months | 'If always treated' versus 'If never treated' |
| FG | 0.178 | 0.057 | 1.70E-03 | 0.067 | 0.290 | 18 months | 'If always treated' versus 'If never treated' |
| BMI | 0.789 | 0.169 | 3.10E-06 | 0.458 | 1.121 | 6 months | 'If always treated' versus 'If never treated' |
| BMI | 0.654 | 0.231 | 4.66E-03 | 0.201 | 1.107 | 9 months | 'If always treated' versus 'If never treated' |
| BMI | 0.903 | 0.198 | 4.89E-06 | 0.516 | 1.291 | 12 months | 'If always treated' versus 'If never treated' |
| BMI | 1.006 | 0.235 | 1.79E-05 | 0.547 | 1.466 | 15 months | 'If always treated' versus 'If never treated' |
| BMI | 0.947 | 0.248 | 1.35E-04 | 0.461 | 1.433 | 18 months | 'If always treated' versus 'If never treated' |
| TC | 0.150 | 0.059 | 1.09E-02 | 0.035 | 0.266 | 6 months | 'If always treated' versus 'If never treated' |
| TC | 0.203 | 0.077 | 8.12E-03 | 0.053 | 0.352 | 9 months | 'If always treated' versus 'If never treated' |
| TC | 0.248 | 0.064 | 9.42E-05 | 0.124 | 0.373 | 12 months | 'If always treated' versus 'If never treated' |
| TC | 0.289 | 0.075 | 1.22E-04 | 0.142 | 0.437 | 15 months | 'If always treated' versus 'If never treated' |
| TC | 0.283 | 0.084 | 7.38E-04 | 0.118 | 0.447 | 18 months | 'If always treated' versus 'If never treated' |
| HDL | -0.074 | 0.007 | 5.75E-25 | -0.088 | -0.060 | 6 months | 'If always treated' versus 'If never treated' |
| HDL | -0.062 | 0.013 | 2.28E-06 | -0.088 | -0.036 | 9 months | 'If always treated' versus 'If never treated' |
| HDL | -0.069 | 0.012 | 4.58E-09 | -0.093 | -0.046 | 12 months | 'If always treated' versus 'If never treated' |
| HDL | -0.069 | 0.015 | 5.47E-06 | -0.099 | -0.039 | 15 months | 'If always treated' versus 'If never treated' |
| HDL | -0.064 | 0.018 | 3.63E-04 | -0.099 | -0.029 | 18 months | 'If always treated' versus 'If never treated' |
| LDL | 0.098 | 0.041 | 1.82E-02 | 0.017 | 0.178 | 6 months | 'If always treated' versus 'If never treated' |
| LDL | 0.145 | 0.062 | 1.86E-02 | 0.024 | 0.266 | 9 months | 'If always treated' versus 'If never treated' |
| LDL | 0.189 | 0.048 | 7.65E-05 | 0.095 | 0.283 | 12 months | 'If always treated' versus 'If never treated' |
| LDL | 0.215 | 0.058 | 2.19E-04 | 0.101 | 0.329 | 15 months | 'If always treated' versus 'If never treated' |
| LDL | 0.205 | 0.062 | 9.58E-04 | 0.083 | 0.327 | 18 months | 'If always treated' versus 'If never treated' |
| Triglycerides | 0.212 | 0.076 | 5.36E-03 | 0.063 | 0.360 | 6 months | 'If always treated' versus 'If never treated' |
| Triglycerides | 0.241 | 0.065 | 2.27E-04 | 0.113 | 0.368 | 9 months | 'If always treated' versus 'If never treated' |
| Triglycerides | 0.028 | 0.261 | 9.15E-01 | -0.484 | 0.540 | 12 months | 'If always treated' versus 'If never treated' |
| Triglycerides | 0.346 | 0.054 | 1.49E-10 | 0.240 | 0.452 | 15 months | 'If always treated' versus 'If never treated' |
| Triglycerides | 0.360 | 0.062 | 6.82E-09 | 0.238 | 0.481 | 18 months | 'If always treated' versus 'If never treated' |

1) The interval between each time-point was 3 months.
2) Definition of treatment: treatment with clozapine or olanzapine. The time-varying confounders included the treatment status of four other drugs, including metformin, atorvastatin, simvastatin, and valproate. Please refer to the main text for details.
3) ATE indicates the difference in the average treatment effect between patients taking clozapine or olanzapine and those taking other SGAs.
4) Abbreviations: TC, total cholesterol; HDL, high-density lipoprotein; LDL, low-density lipoprotein; FG, fasting blood glucose level; BMI, body mass index.

**Table 3.** Average treatment effect (ATE) of SGAs comparing different time of SGA discontinuation with never treated.

| Outcome | ATE | Std.error | P value | CI_0.025 | CI_0.975 | Treatment_<br>order | Never<br>treated | Follow-up length |
| --- | --- | --- | --- | --- | --- | --- | --- | --- |
| FG | 0.002 | 0.048 | 9.59E-01 | -0.091 | 0.096 | abar_1_0_0_0 | abar_0_0_0_0 | 12 |
| FG | 0.047 | 0.048 | 3.28E-01 | -0.047 | 0.142 | abar_1_1_0_0 | abar_0_0_0_0 | 12 |
| FG | -0.033 | 0.049 | 5.00E-01 | -0.129 | 0.063 | abar_1_1_1_0 | abar_0_0_0_0 | 12 |
| FG | 0.090 | 0.040 | **2.35E-02** | 0.012 | 0.169 | abar_1_1_1_1 | abar_0_0_0_0 | 12 |
| BMI | 0.326 | 0.143 | **2.29E-02** | 0.045 | 0.607 | abar_1_0_0_0 | abar_0_0_0_0 | 12 |
| BMI | 0.556 | 0.139 | **6.20E-05** | 0.284 | 0.828 | abar_1_1_0_0 | abar_0_0_0_0 | 12 |
| BMI | 0.350 | 0.146 | **1.65E-02** | 0.064 | 0.635 | abar_1_1_1_0 | abar_0_0_0_0 | 12 |
| BMI | 0.811 | 0.092 | **1.41E-18** | 0.630 | 0.991 | abar_1_1_1_1 | abar_0_0_0_0 | 12 |
| TC | -0.029 | 0.058 | 6.09E-01 | -0.142 | 0.083 | abar_1_0_0_0 | abar_0_0_0_0 | 12 |
| TC | 0.019 | 0.057 | 7.39E-01 | -0.094 | 0.132 | abar_1_1_0_0 | abar_0_0_0_0 | 12 |
| TC | -0.032 | 0.060 | 5.87E-01 | -0.149 | 0.084 | abar_1_1_1_0 | abar_0_0_0_0 | 12 |
| TC | 0.146 | 0.050 | **3.61E-03** | 0.048 | 0.244 | abar_1_1_1_1 | abar_0_0_0_0 | 12 |
| HDL | 0.007 | 0.008 | 3.66E-01 | -0.008 | 0.022 | abar_1_0_0_0 | abar_0_0_0_0 | 12 |
| HDL | -0.004 | 0.008 | 6.36E-01 | -0.020 | 0.012 | abar_1_1_0_0 | abar_0_0_0_0 | 12 |
| HDL | 0.001 | 0.008 | 9.44E-01 | -0.016 | 0.017 | abar_1_1_1_0 | abar_0_0_0_0 | 12 |
| HDL | -0.032 | 0.006 | **2.33E-07** | -0.044 | -0.020 | abar_1_1_1_1 | abar_0_0_0_0 | 12 |
| LDL | 0.009 | 0.046 | 8.43E-01 | -0.082 | 0.100 | abar_1_0_0_0 | abar_0_0_0_0 | 12 |
| LDL | 0.032 | 0.046 | 4.85E-01 | -0.058 | 0.122 | abar_1_1_0_0 | abar_0_0_0_0 | 12 |
| LDL | -0.009 | 0.046 | 8.46E-01 | -0.098 | 0.080 | abar_1_1_1_0 | abar_0_0_0_0 | 12 |
| LDL | 0.121 | 0.040 | **2.62E-03** | 0.042 | 0.200 | abar_1_1_1_1 | abar_0_0_0_0 | 12 |
| Triglycerides | 0.004 | 0.044 | 9.27E-01 | -0.083 | 0.091 | abar_1_0_0_0 | abar_0_0_0_0 | 12 |
| Triglycerides | 0.069 | 0.056 | 2.21E-01 | -0.041 | 0.180 | abar_1_1_0_0 | abar_0_0_0_0 | 12 |
| Triglycerides | 0.023 | 0.054 | 6.78E-01 | -0.084 | 0.129 | abar_1_1_1_0 | abar_0_0_0_0 | 12 |
| Triglycerides | 0.241 | 0.044 | **4.82E-08** | 0.155 | 0.328 | abar_1_1_1_1 | abar_0_0_0_0 | 12 |

- 1) The interval between each time-point was 3 months. - 2) The definition of treatment (7 SGAs): taking any of these SGAs, including clozapine, olanzapine, amisulpride, paliperidone, risperidone, quetiapine, or lurasidone; the definition of time-varying confounders (4 drugs): taking any of these drugs, including metformin, atorvastatin, simvastatin, or valproate. - 3) Treatment is coded as 1 if the patient took SGAs at the observed time point and 0 otherwise. For example, abar\_1\_0\_0\_0 indicates SGA treatment at the 1<sup>st</sup> time-point (3<sup>rd</sup> month) but not afterwards. - 4) Abbreviation: TC, total cholesterol; HDL, high-density lipoprotein; LDL, low-density lipoprotein; FG, fasting blood glucose level; BMI, body mass index

### Clinical implications

Our findings have several clinical implications. The observation that BMI remains elevated after SGA discontinuation is an important finding to clinicians and patients. Our findings suggest that certain metabolic side-effects of SGAs may be relatively long-lasting, even after treatment cessation. Ongoing monitoring of weight/BMI or other obesity indicators, and measures to promote a healthy weight, such as proper diet and exercise, may be beneficial even after SGA discontinuation.

On the other hand, we did not observe any lingering side-effects on other metabolic measures, provided that SGAs have been discontinued for at least 3 months. This suggests that lingering adverse metabolic effects, if present, may be less pronounced for other metabolic measures. However, it should be noted that non-significant results may be due to insufficient power to detect modest differences.

In addition, we observed that with continuation of SGAs, metabolic side-effects increased quickly and peaked at around 12–15 months. These findings may be useful for counseling patients on the naturalistic progression of metabolic side-effects. Clinicians and patients should be particularly aware of the metabolic side-effects emerging in the first 12–15 months of SGA prescription, with possibly more frequent monitoring during this period.

Nevertheless, regardless of the treatment duration, the metabolic outcomes for those on continuous SGA therapy were consistently worse compared to those never on SGAs. This underscores the importance of careful consideration of SGA prescription, and continuous monitoring and management of metabolic health for all patients on these drugs.

Moreover, clozapine and olanzapine appeared to be more strongly linked to greater metabolic side-effects than other SGAs. While stronger metabolic side-effects of these drugs have been reported, we provide further support for these findings using a rigorous *causal* statistical framework which accounts for *time-varying* confounding and treatment status.

### Strengths and limitations

Our study has several notable strengths. First, we employed a longitudinal TMLE framework to quantify the side-effects of SGAs on six metabolic parameters, effectively accounting for the effects of confounders.

Second, our study considered the effects of a *sequence* of treatments (rather than focusing on a single SGA), allowing for changes in SGAs and treatment status during the follow-up period. Our method also accounted for varying follow-up durations at different time-points, which would be challenging to evaluate in randomized controlled trials.

Third, LTMLE is a doubly robust model, meaning that it is consistent if either the outcome model or the treatment model is correctly specified. It contrasts with the traditional methods, which typically rely on a single model.

Fourth, the change in prescriptions was meticulously recorded, and incorporated into the LTMLE model for analysis. Previous studies^6^ have rarely considered changes in medications during follow-ups with the same level of detail. In addition, multiple sensitivity analyses, such as different time intervals and different definitions of treatment status, have supported the robustness of our findings. Also, our study covered a comprehensive range of metabolic outcomes, providing an in-depth understanding of the long-term side-effects of SGAs.

From a methodological perspective, the current study illustrates how LTMLE may be employed to gain clinical insight into an important question, namely SGA’s metabolic side-effects, taking into account the *dynamic* nature of treatment sequences. To our knowledge, very few psychopharmacology studies accounted for treatment sequences, making our study a valuable template for future research in this under-explored area.

However, our study has several limitations. Owing to the limited sample size, we did not analyze the longitudinal effects of each specific SGA but only compared clozapine/olanzapine versus other SGAs. Moreover, non-significant results may be due to inadequate power to detect small effects. In addition, as an observational study, unobserved confounders may be present, although we have employed an advanced statistical approach to address complex and time-varying confounding factors. Lifestyle factors, such as diet, physical activity, and smoking behaviors, were not captured or modeled, which could have influenced the metabolic outcomes.

In conclusion, increasing trends of ATEs of SGAs on BMI, triglycerides, TC, and LDL were observed when the length of treatment increased from 6 to 12/15 months. At 18 months, a decrease in ATEs was observed compared to 12 or 15 months, indicating that the side-effects of SGAs do not continue increasing indefinitely. Lingering adverse effects of SGAs on BMI was observed, although such persistent side-effects were not evident for other metabolic outcomes. Further studies are needed to verify these findings.

## Data Availability

The data that support the findings of this study are available on request from the
corresponding author, HCS. The data are not publicly available due to their containing
information that could compromise the privacy of research participants.

## Notes

### Competing Interest Statement

The authors have declared no competing interest.

### Funding Statement

This work was supported partially by a National Natural Science Foundation China grant (81971706), a National Natural Science Foundation China (NSFC) Young Scientist Grant (31900495), the Lo Kwee Seong Biomedical Research Fund from The Chinese University of Hong Kong and the KIZ-CUHK Joint Laboratory of Bioresources and Molecular Research of Common Diseases, Kunming Institute of Zoology and The Chinese University of Hong Kong, China.

### Author Declarations

Ethical approval has been obtained from the New Territories West Cluster Ethics Committee (approval numbers: NTWC/CREC/823/10 and NTWC/CREC/1293/14) and the Joint Chinese University of Hong Kong-New Territories East Cluster Clinical Research Ethics Committee (approval number: 2016.559). All participants provided written informed consent.

